# No Difference in Risk of Acute Kidney Injury Between Adult Patients Receiving Different Types of Extracorporeal Membrane Oxygenation

**DOI:** 10.1101/2020.03.25.20043950

**Authors:** Zhixiang Mou, Xu Zhang

**Affiliations:** Faculty of Medicine, RWTH Aachen University, Aachen Germany; Department of Nephrology, The First Affiliated Hospital of Dalian Medical University, Dalian China

**Keywords:** AKI, extracorporeal membrane oxygenation, ECMO, ECLS, risk

## Abstract

Acute kidney injury (AKI) has been reported as one of the most common complications in patients receiving extracorporeal membrane oxygenation (ECMO), yet the risk of AKI on different types of ECMO remains unclear. This meta-analysis aimed to compare risk of AKI among adult patients requiring different types of ECMO. Two authors independently performed a literature search using PubMed, Web of Science, and Embase, encompassing publications up until Arpril 20, 2020 (inclusive). The number of AKI patients, non-AKI patients, patients required RRT and patients not required RRT receiving different types of ECMO were derived and analyzed by STATA. The results indicated there was no significant difference in risk of AKI (OR, 1.54; 95% CI: 0.75-3.16; *P*= 0.235) and severe AKI required RRT (OR, 1.0; 95% CI: 0.66-1.5, *P*= 0.994) in patients receiving different types of ECMO. In Conclusion, no difference in risk of AKI and severe AKI required RRT between patients receiving VA ECMO and VV ECMO. More studies are required to support the findings.

## 1. Introduction

Extracorporeal membrane oxygenation (ECMO) is a rescue therapy applied to patients with life-threatening cardiac and/or pulmonary failure and that are unresponsive to conventional therapy.^1^ There has been a rapid increase in the number of ECMO placements across the world during the past few decades.^1, 2^ On the basis of the destination of returned oxygenated blood, ECMO therapy could be subdivided into two basic types: veno-venous (VV) and veno-arterial (VA). VV ECMO drains blood from a central vein and returns the oxygenated blood to another central vein, which results in the venous blood being refreshed by providing gas exchange^3^; however, VA ECMO drains blood from the central vein and pumps the oxygenated blood back to an artery, which involves raising both the arterial pressure and arterial oxygenation.^3^ Our understanding regarding the implications of these two distinct methods for ECMO continue to evolve, but nonetheless it remains a recommended therapy with clear positives, including: resuscitating patients from severe cardiac dysfunction (such as acute cardiogenic shock, cardiac arrest or cardiac failure); acting as a temporary bridge to mechanical assistant device implantations or heart combined/or lung transplantation; post heart or heart-lung transplant failure; any potentially reversible acute respiratory distress syndrome (ARDS)/respiratory failure associated with/without pneumonia (viral or bacterial); a means of providing time to identify other conventional treatments which may effective.^4-6^ Both VV ECMO and VA ECMO can be administered for refractory acute respiratory failure.^7^ The complications of all ECMO technology are similar, and can be divided into complications related to ECMO equipment (oxygenator or pump malfunction, circuit or cannula issues, and so forth) and physiological complications (limb ischemia, stroke, massive bleeding, increased infection or new onset sepsis, haemolysis, severe damage of organs, and so forth). Complications of either category can result in a sub-standard prognosis.^7^ Among all the complications, acute kidney injury (AKI) is the most frequently reported, and appears to be related with poorer prognosis and higher mortality.^8-11^ The causes of AKI which develop during ECMO are multifactorial and extremely complicated, including both patient- and ECMO-related factors.^12^ There are insufficient clinical data available for ascertaining the link between ECMO and AKI. Previously studies have compared the incidence rates and mortality of AKI among patients receiving ECMO, or the differences between reported-only AKI incidences based on different ECMO types via single-center records, which has obvious limitations regarding generalization of findings.^8,11^ There still remains a lack of systematic analysis for the AKI risk between patients receiving different types of ECMO. In addition, the risk of severe AKI associated with renal replacement therapy (RRT) among patients receiving different types of ECMO also remains unclear.

The aim of this meta-analysis is to pool the available published data to compare the risk of AKI among adult patients requiring different types of ECMO.

## 2. Materials and Methods

### 2.1. Search Strategy

The protocol for this meta-analysis has been registered with the International Prospective Register of Systematic Reviews (PROSPERO no. CRD42020178055). A systematic literature review was performed by two authors (ZX.M. and X.Z.) independently through PubMed, Web of Science, and Embase employing the search terms “acute kidney” OR “acute renal” AND “extracorporeal membrane oxygenation” OR “ECMO” OR “extracorporeal life support” OR “ECLS” including publications up until April 20, 2020 (inclusive), to investigate the AKI incidents among adult patients on different types of ECMO. Each study was evaluated for inclusion or exclusion in this analysis (see below). No language or date restrictions were applied. Manual examination for references cited in the analyzed articles was also undertaken. This meta-analysis was conducted and reported according to the guidelines of the Preferred Reporting Items for Systematic Reviews and Meta-Analyses (PRISMA). (http://www.prismastatement.org/).

### 2.2. Study Selection Criteria

In order to fulfil the purpose of this analysis, eligible studies must meet the Population, Interventions, Comparison and Outcomes (PICO) criteria and report the AKI event which occurred in adult patients who underwent ECMO.

All the studies fulfilled the following criteria: original cohort studies provided the data of AKI required/not required RRT among adult patients (age ≥ 16 years) receiving ECMO with consideration of ECMO types as an intervention factor, and potentially eligible studies regardless AKI or RRT definition.

Studies comprising patients with a history of kidney diseases or dialysis required pre-ECMO which cannot be identified after receiving ECMO were excluded; studies which describe “acute kidney disease” without following any AKI-defined guidelines were excluded; studies reporting AKI incidents only during a single type ECMO were excluded; case/case series reports containing < 10 patients were excluded; duplicate articles were excluded; no restrictions on language/year applied in the full text.

### 2.3. Data Extraction and Study Quality

A spreadsheet template (Excel: Microsoft Corporation, Redmond, WA) was built to extract the following data from each included study: the first author, publication year, region, number of patients, percentage of ECMO types, AKI definition, and RRT definition. Some studies required special and very careful review in order to calculate the accuracy of related data.

All included studies were assessed for their quality by the Newcastle-Ottawa Quality Assessment Scale (NOS) comprising three aspects (selection, comparability and outcomes) and eight items.^13^ This enables the researcher to score from 0 to 9, whereby studies with a score ≧ 6 were considered to be of high methodological quality.

### 2.4. AKI and RRT definition

The three widely accepted criteria to define AKI are RIFLE (the Risk of renal failure, Injury to the kidney, Failure of kidney function, Loss of kidney function and End stage kidney disease), AKIN (the Acute Kidney Injury Network), and KDIGO (the Kidney Disease Improving Global Outcomes).^14-15^ The most commonly used RRT modality in ECMO patients is continuous renal replacement therapy (CRRT).^16^ In this meta-analysis, AKI incidents were accepted as long as the original studies had been defined according to at least one of the guidance frameworks cited above; RRT required during ECMO were recognized regardless of implantation modality and KDIGO stage 3.

### 2.5. Statistical Analysis

STATA statistical software (version 14.0: Stata Corp, College Station, TX) was utilized for statistical analysis purposes. In this analysis, random effects models and the Mantel-Haenszel method were applied to analyze the number of AKI patients, non-AKI patients, and patients required/not required RRT during different types of ECMO (dichotomous data); an *I*^2^ test was used to assess heterogeneity due to probability variance among observational studies; the pooled odds ratio (ORs) and their corresponding 95% confidence intervals (CI) were used to evaluate the risk of AKI and the risk of severe AKI required RRT between ECMO types; a *Z* test was used to assess the significance of the pooled ORs and is graphically plotted using forest plots. Statistically significant heterogeneity among studies is defined as χ^2^*P* value< 0.05 or *I*^2^ test > 50%; *Z* test *P* value < 0.05 was considered statistically significant.

## 3. Results

### 3.1 Literature Search and Study Characteristics

The flowchart of the systematic review with selection process and reasons for exclusion is presented in detail in Figure 1. A total of 4,416 records were identified from 3 databases (PubMed, n= 1,445; Web of Science, n= 617; Embase, n= 2,354). After duplicates were excluded (2,000 records), 2,416 articles were screened through titles and abstracts manually for eligibility. Then, in-vitro studies, those focused on pediatric or neonatal populations, animal studies, case reports, conference abstracts and review articles were excluded; 57 remaining articles were full-text reviewed for eligibility. The remaining studies containing < 10 patients or insufficient clinical data were also excluded. Finally, 8 cohort studies involving 1,112 patients which fulfilled the pre-specified criteria were included for the meta-analysis (Table 1). Seven studies (87.5%) with a score ≧ 6 were considered to be of with high quality according to the NOS criteria (Table 2 and Table 3). Five studies (62.5%) reported the outcomes with clearly-defined AKI, according to at least one of the guidelines proposed by RIFLE, AKIN, or KDIGO. Three studies (37.5%) reported RRT required during ECMO regardless established modality.

**Table 1.** Study characteristics

| Authors | Year | Region | Types | No.of patients | AKI definition | RRT definition |
| --- | --- | --- | --- | --- | --- | --- |
| Aubron et al [17] | 2013 | Australia | VA-ECMO 105 (67%)<br>VV-ECMO 53 (33%) | 158 | N/A | Not mentioned |
| Schmidt et al [18] | 2014 | Australia | VA-ECMO 115 (45%)<br>VV-ECMO 57 (55%) | 172 | N/A | CRRT |
| Lee et al [19] | 2015 | Korea | VA-ECMO 230 (71%)<br>VV-ECMO 92 (29%) | 322 | KDIGO | KDIGO stage 3 |
| Antonucci et al [20] | 2016 | Belgium | VA-ECMO 79 (59%)<br>VV-ECMO 56 (41%) | 135 | AKIN | CRRT |
| Chang et al [21] | 2017 | China | VA-ECMO 46 (94%)<br>VV-ECMO 25 ( 6%) | 71 | AKIN | N/A |
| Silva et al [22] | 2017 | Portugal | VA-ECMO 29 (94%)<br>VV-ECMO 19 ( 6%) | 48 | AKIN | Not mentioned |
| Liao et al [23] | 2018 | China | VA-ECMO 159 (94%)<br>VV-ECMO 11 ( 6%) | 170 | KDIGO | N/A |
| Wu et al [24] | 2018 | Taiwan | VA-ECMO 14 (39%)<br>VV-ECMO 22 (61%) | 36 | Not mentioned | CRRT |

**Table 2.**
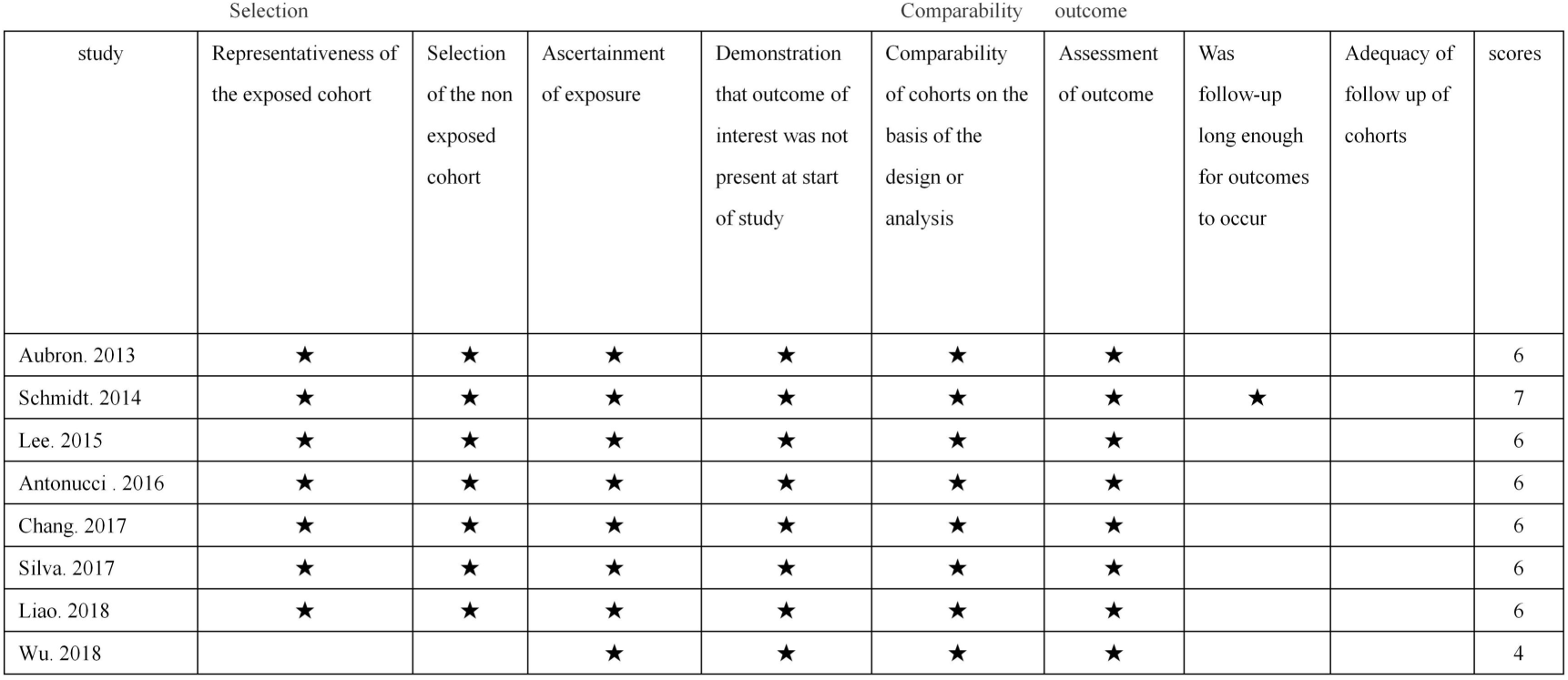
Newcastle-Ottawa Scale for assessing the quality of cohort studies

**Figure. 1.**
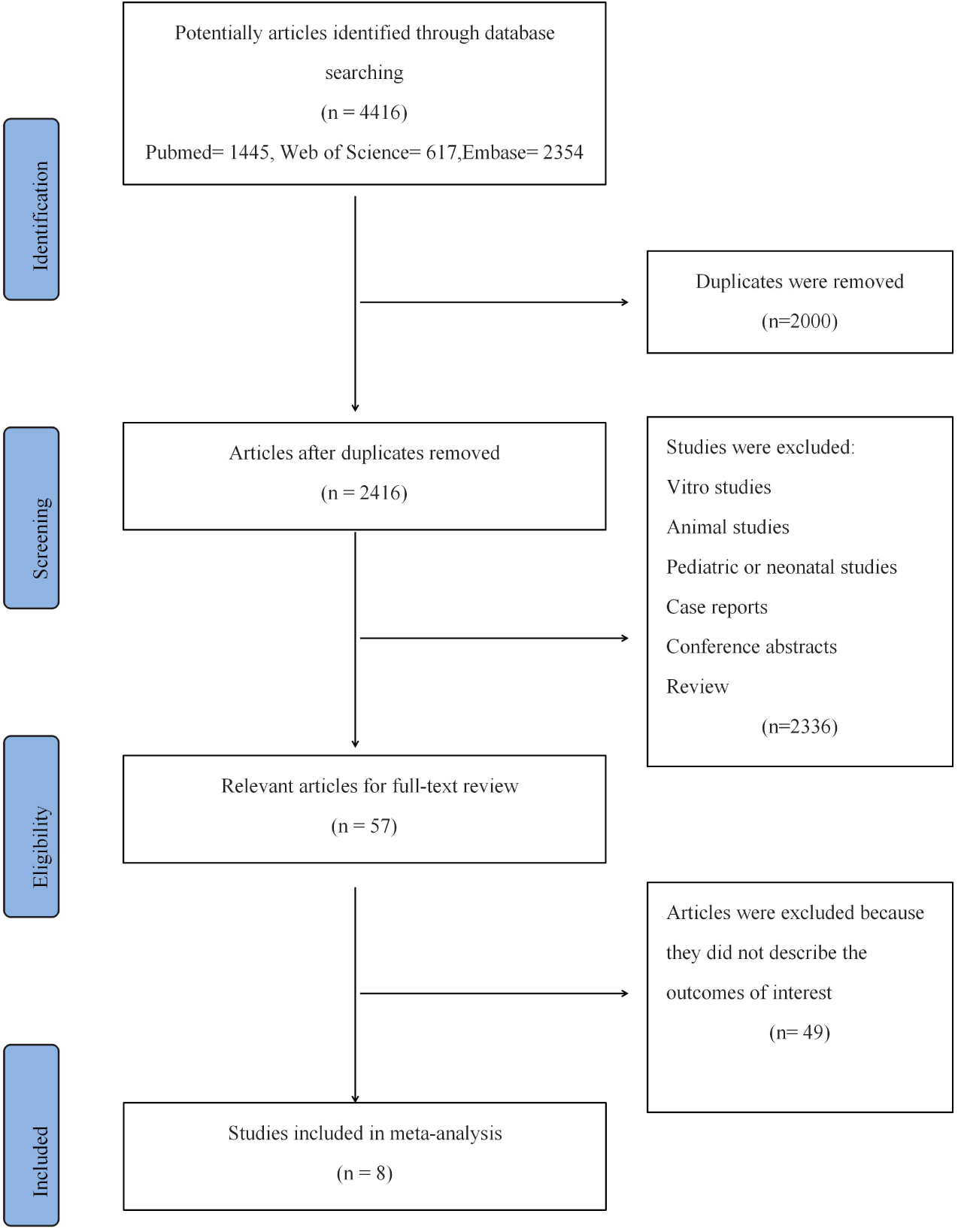
The flowchart for the systematic review.

### 3.2 Patient Characteristics

In this study, 1,112 patients receiving ECMO treatment between January 2005 and October 2016 were identified as being eligible for analysis. Of these patients, 69.9% (n = 777) received VA ECMO, 30.1% (n = 335) received VV ECMO; 50.2% (n = 390) patients developed AKI during VA ECMO, 42.4% (n = 142) patients developed AKI during VV ECMO; 34.9% (n = 271) patients need RRT due to AKI during VA ECMO, and 42.1% (n = 141) patients need RRT due to AKI during VV ECMO.

### 3.3. Risk of AKI in Patients Receiving Different Types of ECMO

Overall, the AKI risk of the patient group receiving VA ECMO was estimated as being 1.54 times higher (OR, 1.54; *I*^2^ = 64.5%, χ^2^*P* = 0.024) than the VV ECMO group (Figure 2); unexpectedly, there was no significant difference (95% CI: 0.75-3.16; *P* = 0.235). Subgroup analyses were performed according to different AKI-defined criteria. Consistent with the overall analysis, the AKI risk of patients the group receiving VA ECMO was estimated to be 2.14 times higher (OR, 2.14; 95% CI: 1.15-3.97, *I*^2^ = 0.0%, χ^2^*P* = 0.431, *P* = 0.016; see Figure 2) in the AKIN subgroup analysis; While the AKI risk of the patient group receiving VA ECMO was estimated to be 0.75 times lower (OR, 0.75; 95% CI: 0.42-1.34, *I*^2^ = 0.0%, χ^2^*P* = 0.785, *P* = 0.331; see Figure 2) in the KDIGO subgroup analysis. The results of subgroup analyses indicated that significant heterogeneity existed perhaps due to inconsistent AKI definitions.

**Figure 2.**
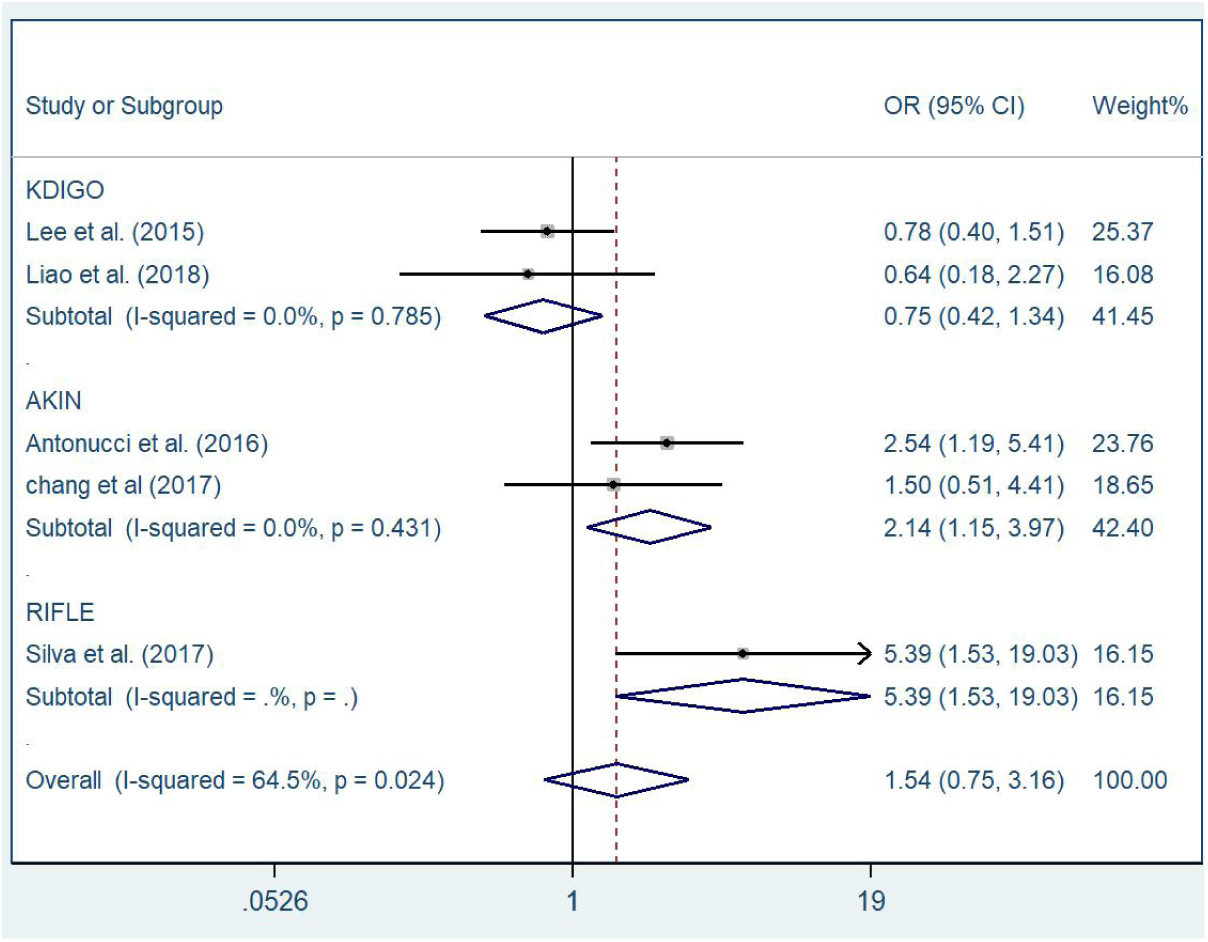
Risk of AKI in Patients Receiving Different Types of ECMO.

### 3.4 Risk of Severe AKI Required RRT in Patients Receiving Different Types of ECMO

The following meta-analysis revealed that there was no no significant difference in risk of severe AKI required RRT in patients receiving different types of ECMO (OR, 1.0; 95% CI: 0.66-1.5, *P* = 0.994; see Figure 3), and significant heterogeneity was not observed (*I*^2^ = 38.3%, χ^2^*P* = 0.15) among the eligible studies.

**Figure 3.**
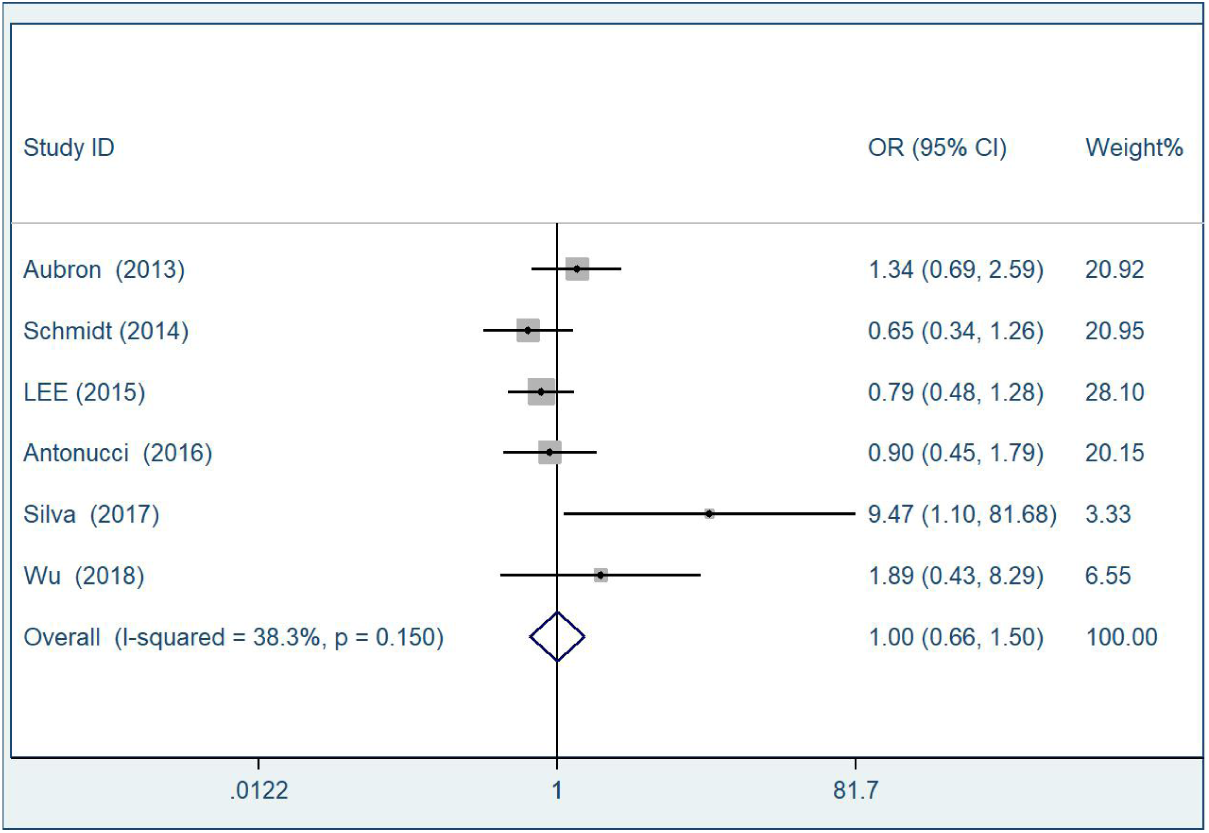
Risk of Severe AKI Required RRT in Patients Receiving Different Types of ECMO.

## 4 DISCUSSION

The population-based meta-analysis herein provides evidence that there is no difference in the risk of AKI between patients receiving VA ECMO and VV ECMO; there is no difference in the risk of severe AKI requiring RRT in patients receiving VA ECMO and VV ECMO.

According to the international voluntary registry Extracorporeal Life Support Organization (ELSO), the number of ECMO incidents has proliferated rapidly, as well as the number of ECMO centers has increased worldwide in the last 40 years^25, 26^ due to improved indication capabilities and technology improvements.^27^ ECMO is not a routine therapy, on account of its complicated techniques and the significant demands on resource. Although new ECMO treatment guidelines are developed in order to facilitate implementation with the highest benefit-to-risk and benefit-to-cost ratios,^28^ patients who receive the treatment still exhibit a high mortality rate and an increased chance of suffering from various complications.^29-31^ The incidence of complications that impact long-term outcomes is varied and depends on the types of patients, primary disease progression, and evolving indications.^4^ AKI is one of the most common complications associated with ECMO, and patients who develop AKI during ECMO would face poorer long-term outcomes and survival rates compared to patients without ECMO-related AKI.^8-11^

The types of ECMO selected by physicians is determined by the types of organs requiring support, the types of patients treated, and the desired outcome.^4^ Most studies^21,22,32^ have considered the incidence of AKI among patients requiring VA ECMO as being higher than those requiring VV ECMO for a long time. VA ECMO is performed for cardiocirculatory support with or without respiratory failure,^33^ simultaneously providing oxygen delivery and cardiac support. Thus, the working cardiac output is a mixture of non-pulsatile arterial flow from the ECMO pump and pulsatile arterial flow from the patient’s native cardiac output. On the other hand, VV ECMO is applied to patients with severe, isolated, and refractory respiratory failures.^33^ It provides oxygenated and decarboxylated blood, retains the native pulsatile cardiac output thus the impact of alterations upon hemodynamics and renal perfusion may be less.^6^ Several recent studies have shown that pulsatile flow may provide more beneficial and protective effects on renal microcirculation and perfusion than non-pulsatile flow,^32, 34, 35^ and these findings are consistent with the differences identified between the working flows of different types of ECMO. Considering these indications, the beneficial effects of VV ECMO may be related to the early correction of blood gas disturbances, enhanced systemic oxygenation and reduced oxygen consumption (which may improve the renal metabolism)^36^ whereas VA ECMO restores adequate end-organ perfusion during low cardiac output in myocardial infarction, end-stage or refractory heart failure, worsening congestion, and cardiogenic shock, which is considered a trigger for AKI (type-1 cardiorenal syndrome).^37^ In addition, pulmonary hypertension and increased right atrial pressures have been identified as risk factors for worsening renal function in patients with heart failure,^38,39^ which is one of the obvious indications for VA-ECMO. When VA-ECMO is established for severe cardiac failure, a low ejection fraction can lead to left ventricular over-distension and worsening right-sided heart failure, causing AKI due to congestion.^40^ A comprehensive review of the different types of ECMO is beyond the scope of this paper, but these fundamental differences between VA and VV ECMO appear to be significant implications, insofar that the risk of AKI in VA ECMO patients seems to be higher than VV ECMO patients.

Unlike evidence of increased mortality in ECMO patients suffering from AKI,^31^ the pathophysiological mechanisms are complex, multifactorial, synergistic and remain poorly understood. The factors inducing AKI during ECMO can be divided into patient-related and ECMO-related, as well as being intertwined with one another. A pathologic dysregulated host response with inflammatory cytokines release due to the primary disease process may induce glycocalyx alteration and microcirculation dysfunction, simultaneously combining with the blood/air interface and exposure of non-self ECMO membrane, would initiate and amplify renal systemic inflammatory and renal injury.^41^ In addition, haemodynamic alterations (such as limited pulsatile, renal hypoperfusion from damaged cardiac function, diuretic/vasopressor therapy, frequently intravenous fluid, high PEEP, variations of ECMO circuit pressures), hormonal issues (renin-angiotensin-aldosterone dysregulation), and circuit-related accidents (embolism, hemolysis, ischemia-reperfusion injury) would act as influential factors and may cause pre-renal AKI to develop to intrinsic AKI, as well as cortical necrosis, finally result in renal dysfunction.^6; 41-44^ The major pathophysiology mechanism of AKI is shown in Figure 4, where the overlapping shaded area can be understood to be the potentially contribution of AKI-related factors. As the authentic accurate pathophysiological mechanism of ECMO related-AKI is not completely clarified, the size of every shaded area is as yet impossible to define. The worst situation is that one key factor maybe powerful enough to cause AKI in critically ill patients during ECMO, although VA ECMO seems to provide a greater contribution.

**Figure 4.**
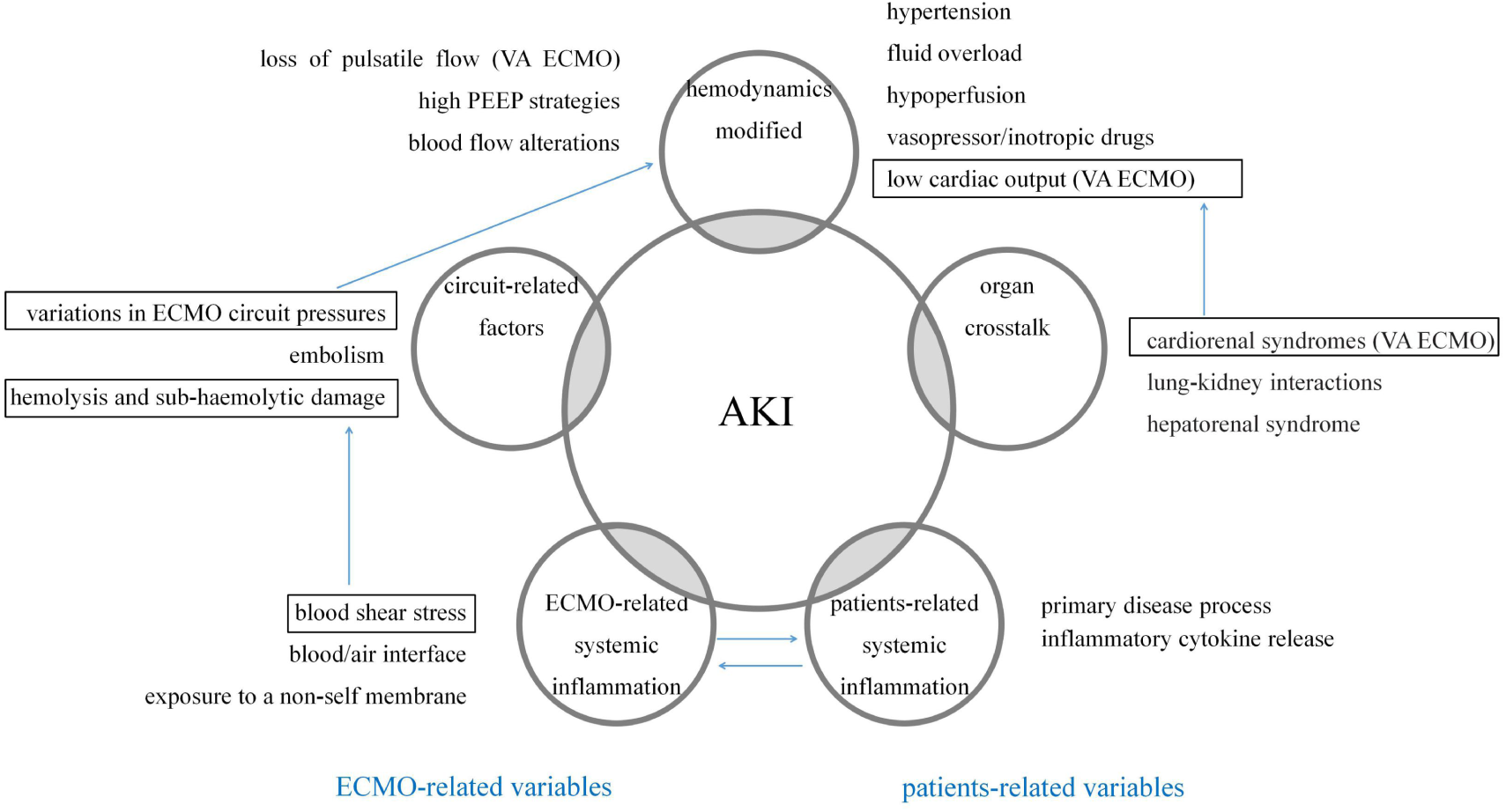
Potential pathophysiological mechanisms of ECMO related AKI.

Regarding conventional treatments, some interventions and approaches are often executed pre-ECMO, with the inevitable sacrifice of renal function, as the physicians try to delay or prevent the ECMO initiation, but also while it is unclear whether the placement of ECMO protects or exacerbates this consequence.^41^ In other words, AKI might be considered an inevitable clinical syndrome in seriously ill patients pre-ECMO, as over 50% of those would develop stage 1 AKI at some point during their ICU course,^45^ particularly in the setting of chronic conditions and multiple pre-existing comorbidities.^40^ According to a recently published viewpoint “organ crosstalk” (cardio-renal syndrome, lung-kidney interactions, hepatorenal syndrome), renal dysfunction can perhaps always act as secondarily accident, involved in other organ failures and interactions, instead of comprising the initiating event,^40^ no matter what type of ECMO implantation is applied.

RRT as a concomitant treatment with ECMO has been routinely used for kidney support over several decades in various ICUs worldwide. The most commonly modality is CRRT. Although the reasons for initiating RRT may vary (fluid overload, acidosis, electrolyte imbalance, or inadequate renal clearance, to name but a few), an overall higher mortality among ECMO patients required RRT than those who required ECMO alone has already been proven.^46,47^ Due to the complicated primary diseases and our continually evolving understanding of ECMO, the intention of this research is to play a role in preventing and identifying as early as possible the patients at potential risk of ECMO-related AKI when faced the option of ECMO types. However, given the findings that no significant differences in the risk of AKI and severe AKI required RRT between patients receiving VA ECMO and VV ECMO in fact, this now seems unnecessary.

The study has several limitations. First, influence analysis and the potential for publication bias analysis cannot be reasonably or effectively performed due to the limited number of original studies (< 10). Second, there is statistical heterogeneity in the meta-analysis for risk of AKI. Potential origins for heterogeneity may lie in the definitions of AKI. The subgroup analysis performed according to the AKI definition established herein, the results show there is no heterogeneity in the AKIN and KDIGO subgroups. The timing of onset of AKI by any definition is indeed difficult to establish, which is why some study^41^ consider that the three traditional definitions are limited and suffer from insufficient accuracy by which to define ECMO-related AKI; this contention is also beyond the scope of the analysis, but has been addressed in other papers.^41^ Finally, although most of the patients developed ECMO-related AKI required RRT within 48 hours of ECMO insertion,^48^ long-term ECMO has been reported as an independent risk factor of AKI.^49^ Since the clinical data based on publications collected herein is limited in terms of availability, extra sub-analyses could not performed (time spent, supporting organs, blood flow rate, and others). Incidentally, further studies are still needed to support the conclusion and demonstrate more associations of AKI among the patients who have undergone ECMO.

## 5. Conclusions

In conclusion, there is no difference in the risk of AKI and severe AKI required RRT between patients who have received VA ECMO and VV ECMO. More studies are needed in order to support or refute the findings and focus on the systematic clinical characteristics to predict and detect AKI associations among ECMO patients.

## Data Availability

All data generated or analyzed during this study are included in this article.

## References

1. Karagiannidis C, Brodie D, Strassmann S, et al. Extracorporeal membrane oxygenation: evolving epidemiology and mortality. Intensive Care Med. 2016;42:889–896.

2. Mao J, Paul S, Sedrakyan A. The evolving use of ECMO: The impact of the CESAR trial. Int J Surg. 2016;35:95–9.

3. Kulkarni T, Sharma NS, Diaz-Guzman E. Extracorporeal membrane oxygenation in adults: a practical guide for internists. Cleve Clin J Med. 2016;83:373–384

4. Butt W, Maclaren G. Extracorporeal membrane oxygenation. F1000Prime Rep. 2013;5:55.

5. Brodie D, Bacchetta M. Extracorporeal membrane oxygenation for ARDS in adults. N Engl J Med. 2011;365:1905–14.

6. Chen YC, Tsai FC, Fang JT, et al. Acute kidney injury in adults receiving extracorporeal membrane oxygenation. J Formos Med Assoc. 2014;113:778–85.

7. Guglin M, Zucker MJ, Bazan VM, et al. Venoarterial ECMO for Adults: JAC C Scientific Expert Panel. J Am Coll Cardiol. 2019;73:698–716.

8. Tsai TY, Chien H, Tsai FC, et al. Comparison of RIFLE, AKIN, and KDIGO classifications for assessing prognosis of patients on extracorporeal membrane oxygenation. J Formos Med Assoc. 2017;116:844–851.

9. Soliman IW, Frencken JF, Peelen LM, et al. The predictive value of early acute kidney injury for long-term survival and quality of life of critically ill patients. Crit Care. 2016;20:242.

10. Ma P, Zhang Z, Song T, et al. Combining ECMO with IABP for the treatment of critically Ill adult heart failure patients. Heart Lung Circ. 2014;23:363–8.

11. Schmidt M, Bailey M, Kelly J, et al. Impact of fluid balance on outcome of adult patients treated with extracorporeal membrane oxygenation. Intensive Care Med. 2014;40:1256–66.

12. Villa G, Katz N, Ronco C. Extracorporeal Membrane Oxygenation and the Kidney. Cardiorenal Med. 2015;6:50–60.

13. Claudio Luchini, Brendon Stubbs, Marco Solmi, et al. Assessing the quality of studies in meta-analyses: Advantages and limitations of the Newcastle Ottawa Scale. World J Meta-Anal. 2017;5:80–84.

14. Bellomo R, Ronco C, Kellum JA, et al. Acute renal failure e definition, outcome measures, animal models, fluid therapy and information technology needs: the Second International Consensus Conference of the Acute Dialysis Quality Initiative (ADQI) group. Crit Care. 2004;8(4):R204–12.

15. Mehta RL, Kellum JA, Shah SV, et al. Acute Kidney Injury Network: report of an initiative to improve outcomes in acute kidney injury. Crit Care. 2007;11(2):R31, 2007.

16. Ostermann M, Connor M Jr, Kashani K. Continuous renal replacement therapy during extracorporeal membrane oxygenation: why, when and how? Curr Opin Crit Care. 2018;24(6):493–503.

17. Aubron C, Cheng AC, Pilcher D, et al. Factors associated with outcomes of patients on extracorporeal membrane oxygenation support: a 5-year cohort study. Crit Care. 2013;17:R73.

18. Schmidt M, Bailey M, Kelly J, et al. Impact of fluid balance on outcome of adult patients treated with extracorporeal membrane oxygenation. Intensive Care Med. 2014;40(9):1256–66.

19. Lee SW, Yu MY, Lee H, et al. Risk Factors for Acute Kidney Injury and In Hospital Mortality in Patients Receiving Extracorporeal Membrane Oxygenation. PLoS One. 2015;10:e0140674.

20. Antonucci E, Lamanna I, Fagnoul D, et al. The Impact of Renal Failure and Renal Replacement Therapy on Outcome During Extracorporeal Membrane Oxygenation Therapy. Artif Organs. 2016;40:746–54.

21. Xin Chang, Zhen Guo, Lingfeng Xu, et al. Acute kidney injury in patients receiving ECMO: risk factors and outcomes. Int J Clin Exp Med. 2017;10(12):16663–9.

22. Passos Silva M, Caeiro D, Fernandes P, et al. Extracorporeal membrane oxygenation in circulatory and respiratory failure - A single-center experience. Rev Port Cardiol. 2017;36:833–842.

23. Liao X, Cheng Z, Wang L, et al. Analysis of the risk factors of acute kidney injury in patients receiving extracorporeal membrane oxygenation. Clin Nephrol. 2018;90:270–275.

24. Wu MY, Chou PL, Wu TI, et al. Predictors of hospital mortality in adult trauma patients receiving extracorporeal membrane oxygenation for advanced life support: a retrospective cohort study. Scand J Trauma Resusc Emerg Med. 2018;26:14.

25. Thiagarajan RR, Barbaro RP, Rycus PT, et al. Extracorporeal Life Support Organization Registry International Report 2016. ASAIO J. 2017;63:60–67.

26. Sauer CM, Yuh DD, Bonde P. Extracorporeal membrane oxygenation use has increased by 433% in adults in the United States from 2006 to 2011. ASAIO J. 2015;61:31–6.

27. Shekar K, Mullany DV, Thomson B, et al. Extracorporeal life support devices and strategies for management of acute cardiorespiratory failure in adult patients: a comprehensive review. Crit Care. 2014;18:219.

28. Rosenberg AA, Haft JW, Bartlett R, et al. Prolonged duration ECMO for ARDS: Futility,native lung recovery, or transplantation? ASAIO J. 2013;59:642–50.

29. Gray BW, Haft JW, Hirsch JC, et al. Extracorporeal life support: experience with 2,000 patients. ASAIO J. 2015;61:2–7.

30. Zangrillo A, Landoni G, Biondi-Zoccai G, et al. A meta-analysis of complications and mortality of extracorporeal membrane oxygenation. Crit Care Resusc. 2013;15:172–8.

31. Tsai CW, Lin YF, Wu VC, et al. SAPS 3 at dialysis commencement is predictive of hospital mortality in patients supported by extracorporeal membrane oxygenation and acute dialysis. Eur J Cardiothorac Surg. 2008;34:1158–64.

32. Thongprayoon C, Cheungpasitporn W, Lertjitbanjong P, et al. Incidence and Impact of Acute Kidney Injury in Patients Receiving Extracorporeal Membrane Oxygenation: A Meta-Analysis. J Clin Med. 2019;8:E981.

33. Adademir T, Ak K, Aljodi M, et al. The effects of pulsatile cardiopulmonary bypass on acute kidney injury. Int J Artif Organs. 2012;35:511–9.

34. Abu-Omar Y, Ratnatunga C. Cardiopulmonary bypass and renal injury. Perfusion. 2006;21:209–13.

35. Santana-Santos E, Marcusso ME, Rodrigues AO, et al. Strategies for prevention of acute kidney injury in cardiac surgery: an integrative review. Rev Bras Ter Intensiva. 2014;26:183–92.

36. Lyu L, Long C, Hei F, et al. plasma free hemoglobin is a predictor of acute renal failure during adult venous-arterial extracorporeal membrane oxygenation support. J Cardiothorac Vasc Anesth. 2016;30:891–895.

37. Ronco C, Bellasi A, Di Lullo L. Cardiorenal Syndrome: An Overview. Adv Chronic Kidney Dis. 2018;25(5):382–390.

38. L.M. Mielniczuk, G. Chandy, D. Stewart, et al. Worsening renal function and prognosis in pulmonary hypertension patients hospitalized for right heart failure, Congest. Heart Fail. 2012;18(3):151–7.

39. Ivey-Miranda JB, Posada-Martínez EL, Almeida-Gutiérrez E, et al. Right atrial pressure predicts worsening renal function in patients with acute right ventricular myocardial infarction. Int J Cardiol. 2018;264:25–27.

40. Husain-Syed F, Ricci Z, Brodie D, et al. Extracorporeal organ support (ECOS) in critical illness and acute kidney injury: from native to artificial organ crosstalk. Intensive Care Med. 2018;44(9):1447–1459.

41. Kilburn DJ, Shekar K, Fraser JF. The Complex Relationship of Extracorporeal Membrane Oxygenation and Acute Kidney Injury: Causation or Association? Biomed Res Int. 2016;2016:1094296.

42. Hamdi T, Palmer BF. Review of Extracorporeal Membrane Oxygenation and Dialysis-Based Liver Support Devices for the Use of Nephrologists. Am J Nephrol. 2017;46:139–149.

43. McDonald CI, Fraser JF, Coombes JS, et al. Oxidative stress during extracorporeal circulation. Eur J Cardiothorac Surg. 2014;46:937–43.

44. Ikeda M, Prachasilchai W, Burne-Taney MJ, et al. Ischemic acute tubular necrosis models and drug discovery: A focus on cellular inflammation. Drug Discov Today. 2006;11:364–70.

45. Hoste EA, Bagshaw SM, Bellomo R, et al. Epidemiology of acute kidney injury in critically ill patients: the multinational AKI-EPI study. Intensive Care Med. 2015; 41(8):1411–1423.

46. Chen SW, Lu YA, Lee CC, et al. Long-term outcomes after extracorporeal membrane oxygenation in patients with dialysis-requiring acute kidney injury: A cohort study. PLoS One. 2019;14:e0212352.

47. Askenazi DJ, Selewski DT, Paden ML, et al. Renal replacement therapy in critically ill patients receiving extracorporeal membrane oxygenation. Clin J Am Soc Nephrol. 2012;7:1328–36.

48. Kielstein JT, Heiden AM, Beutel G, et al. Renal function and survival in 200 patients undergoing ECMO therapy. Nephrol Dial Transplant. 2013;28:86–90.

49. Reeb J, Olland A, Renaud S, et al. Vascular access for extracorporeal life support: tips and tricks. J Thorac Dis. 2016;8:S353–63.

